# Predictive accuracy of computer-aided versions of the on-admission National Early Warning Score in estimating the risk of COVID-19 for unplanned admission to hospital: a retrospective development and validation study

**DOI:** 10.1101/2020.11.30.20241257

**Authors:** Muhammad Faisal, Mohammed A Mohammed, Donald Richardson, Ewout W. Steyerberg, Massimo Fiori, Kevin Beatson

## Abstract

**Objectives:** To consider the potential of the National Early Warning Score (NEWS2) for COVID-19 risk prediction on unplanned admission to hospital.

**Design:** Logistic regression model development and validation study using a cohort of unplanned emergency medical admission to hospital.

**Setting:** York Hospital (YH) as model development dataset and Scarborough Hospital (SH) as model validation dataset.

**Participants:** Unplanned adult medical admissions discharged over 3 months (11 March 2020 to 13 June 2020) from two hospitals (YH for model development; SH for external model validation) based on admission NEWS2 electronically recorded within ±24 hours of admission. We used logistic regression modelling to predict the risk of COVID-19 using NEWS2 (Model M0’) versus enhanced cNEWS models which included age + sex (model M1’) + subcomponents (including diastolic blood pressure + oxygen flow rate + oxygen scale) of NEWS2 (model M2’). The ICD-10 code ‘U071’ was used to identify COVID-19 admissions. Model performance was evaluated according to discrimination (c statistic), calibration (graphically), and clinical usefulness at NEWS2 ≥5.

**Results:** The prevalence of COVID-19 was higher in SH (11.0%=277/2520) than YH (8.7%=343/3924) with higher index NEWS2 (3.2 vs 2.8) but similar in-hospital mortality (8.4% vs 8.2%). The c-statistics for predicting COVID-19 for cNEWS models (M1’,M2’) was substantially better than NEWS2 alone (M0’) in development (M2’: 0.78 (95%CI 0.75-0.80) vs M0’ 0.71 (95%CI 0.68-0.74)) and validation datasets (M2’: 0.72 (95%CI 0.69-0.75) vs M0’ 0.65 (95%CI 0.61-0.68)). Model M2’ had better calibration than Model M0’ with improved sensitivity (M2’: 57% (95%CI 51%-63%) vs M0’ 44% (95%CI 38%-50%)) and similar specificity (M2’: 76% (95%CI 74%-78%) vs M0’ 75% (95%CI 73%-77%)) for validation dataset at NEWS2≥5.

**Conclusions:** Model M2’ is reasonably accurate for predicting the on-admission risk of COVID-19. It may be clinically useful for an early warning system at the time of admission especially to triage large numbers of unplanned hospital admissions.

## Introduction

The novel coronavirus SARS-19 produces the newly identified disease ‘COVID-19’ in patients with symptoms (Coronaviridae Study Group of the International Committee on Taxonomy of Viruses(1)) which was declared as a pandemic on 11-March-2020 that has challenged health care systems worldwide. COVID-19 patients admitted to hospital can develop severe disease with life threatening respiratory and/or multi-organ failure (2, 3) with a high risk of mortality in part due to the lack of an effective treatment (bar supportive care) for the underlying disease. The appropriate early assessment and management of patients with COVID-19 is important in ensuring high-quality care including isolation, escalation to critical care or palliative care. Early assessment of the risk of COVID-19 is crucial to this process. Presently this involves clinical judgment based on the patients presenting history, signs and symptoms and viral nucleic acid testing can have a 24-hour turnaround time (4).

We posit that vital signs, which form the basis of Early Waning Scores (EWS) may be useful in supporting the clinical decision-making process before swab test results are available. EWS, which are widely used in hospitals worldwide, and in the National Health Service (NHS) hospitals in England, the patient’s vital signs are monitored and summarised into a National Early Warning Score (NEWS)(5). NEWS offers a standardised approach to assessing acute illness and is derived from seven physiological variables or vital signs – respiration rate, oxygen saturations, any supplemental oxygen, temperature, systolic blood pressure, heart rate and level of consciousness (Alert (A), Voice (V), Pain (P), Unresponsive (U)) – which are routinely collected by nursing staff as an integral part of the process of care.

NEWS was launched by the Royal College of Physicians in 2012 (5) and has gained widespread interest from across the world, including Europe, India, the USA (and the US Navy)(6). In December 2017, an update to NEWS (NEWS2) was published (6) that extends the level of consciousness from AVPU to ACVPU, where C represents new confusion or delirium and is allocated 3 points (the maximum for a single variable). NEWS2 also offers two scales for oxygen saturation (scale 1 and scale 2) which accommodates patients with hypercapnic respiratory failure who have clinically recommended oxygen saturation of 88–92%.

Whilst hospitals continue to use NEWS2 during the COVID-19 pandemic, we determine the extent to which NEWS2 can be used predict the risk of COVID-19 by computer enhanced NEWS2 models. This approach is clinically useful because it places no additional data collection burden on staff whilst having the potential of providing an early indication of COVID-19 risk before findings of a swab test are reported - thus supporting early triage of COVID-19 and non-COVID-19 patients.

## Methods

### Setting & data

Our cohorts of emergency medical admissions are from two acute hospitals which are approximately 65 kilometres apart in the Yorkshire & Humberside region of England – Scarborough hospital (n∼300 beds) and York Hospital (YH) (n∼700 beds), managed by York Teaching Hospitals NHS Foundation Trust. We selected these hospitals because they had electronic NEWS2, which are collected as part of the patient’s process of care and were agreeable to the study. Since NEWS2 extends NEWS, we use the same dataset to develop and validate NEWS and NEWS2 models, especially as NEWS is still in widespread use.

We considered all consecutive adult (age≥18 years) non-elective or emergency medical admissions discharged during 3 months (11 March 2020 to 13 June 2020), with electronic NEWS2. For each emergency admission, we obtained a pseudonymised patient identifier, patient’s age (years), gender (male/female), discharge status (alive/dead), admission and discharge date and time, diagnoses codes based on the 10th revision of the International Statistical Classification of Diseases (ICD-10), NEWS2 (including its subcomponents respiratory rate, temperature, systolic pressure, pulse rate, oxygen saturation, oxygen supplementation, oxygen scales 1 & 2, and alertness including confusion). The diastolic blood pressure was recorded at the same time as systolic blood pressure. Historically, diastolic blood pressure has always been a routinely collected physiological variable on vital sign charts and is still collected where electronic observations are in place. NEWS2 produces integer values that range from 0 (indicating the lowest severity of illness) to 20 (the maximum NEWS2 value possible) (see Table S1 and S2 in supplementary material). The index NEWS2 was defined as the first electronically recorded NEWS2 within ±24 hours of the admission time. We excluded records where the index NEWS2 was not within ±24 hours or was missing/not recorded at all (see Table 1). The ICD-10 code ‘U071’ was used to identify records with COVID-19. We searched primary and secondary ICD-10 codes for ‘U071’ for identifying COVID-19.

**Table 1.** Characteristics of emergency medical admissions in development and validation datasets.

| Characteristic | Development dataset (YH) | Validation dataset (SH) | Degree of freedom (df) | p-value |
| --- | --- | --- | --- | --- |
| N | 3924 | 2520 |  | - |
| Male (%) | 2010 (51.2) | 1247 (49.5) | 1 | 0.181 |
| Mean Age [years] (SD) | 67.4 (18.7) | 69.6 (18.9) | 5320 | <0.001 |
| Median Length of Stay (days) (IQR) | 3.0 (5.8) | 3.7 (6.1) | - | <0.001 |
| COVID-19 (%) | 343 (8.7) | 277 (11.0) | 1 | 0.003 |
| **Mortality** |  |  |  |  |
| Mortality with-in 24 hours (%) | 30 (0.8) | 32 (1.3) | 1 | 0.058 |
| Mortality with-in 48 hours (%) | 61 (1.6) | 48 (1.9) | 1 | 0.335 |
| Mortality with-in 72 hours (%) | 96 (2.4) | 68 (2.7) | 1 | 0.585 |
| In-hospital Mortality | 323 (8.2) | 212 (8.4) | 1 | 0.833 |
| Mean NEWS (SD) | 2.5 (2.3) | 2.8 (2.4) | 5201 | <0.001 |
| Mean NEWS2 (SD) | 2.8 (2.8) | 3.2 (2.8) | 5446 | <0.001 |
| **Vital Signs** |  |  |  |  |
| Mean Respiratory rate [breaths per minute] (SD) | 19.8 (5.1) | 20.7 (5.6) | 5027 | <0.001 |
| Mean Temperature [°C] (SD) | 36.4 (0.9) | 36.3 (1) | 4817 | 0.001 |
| Mean Systolic pressure [mmHg] (SD) | 141.8 (29.2) | 142 (28.5) | 5455 | 0.839 |
| Mean Diastolic pressure [mmHg] (SD) | 79.2 (16.5) | 79 (17.3) | 5193 | 0.545 |
| Mean Pulse rate [beats per minute] (SD) | 89.1 (22.3) | 88.5 (22.1) | 5406 | 0.336 |
| Mean Oxygen saturation (SD) | 96.3 (3.1) | 96.1 (3.2) | 5182 | 0.059 |
| Oxygen supplementation (%) | 512 (13) | 362 (14.4) | 1 | 0.142 |
| Mean oxygen flow rate (units) (SD) | 7.1 (5.7) | 6.1 (5.3) | 811 | 0.007 |
| Oxygen scale 2 (yes) (%) | 240 (6.1) | 163 (6.5) | 1 | 0.605 |
| **Alertness** |  |  |  |  |
| Alert (%) | 3510 (89.4) | 2243 (89) | 5 | 0.010 |
| Baseline confusion (%) | 27 (0.7) | 23 (0.9) |  |  |
| New confusion (%) | 61 (1.6) | 40 (1.6) |  |  |
| Pain (%) | 32 (0.8) | 17 (0.7) |  |  |
| Voice (%) | 151 (3.8) | 134 (5.3) |  |  |
| Unconscious (%) | 143 (3.6) | 63 (2.5) |  |  |

### Statistical Modelling

We began with exploratory analyses including box plots that showed the relationship between covariates and risk of COVID-19 and line plots showed the relationship between age, vital signs, NEWS2 and risk of COVID-19. We developed three logistic regression models for each NEWS and NEWS2 separately predicting the risk of COVID-19. The NEWS models (M0, M1, M2) use the index or first recorded NEWS within ±24hours of admission. Model M0 uses NEWS alone; Model M1 extends M0 with age and sex and Model M2 extends M1 with all the subcomponents of NEWS plus diastolic blood pressure. Equivalent models (M0’, M1’, M2’) using NEWS2 were also developed but they include oxygen flow rate as a continuous covariate and scale 1/scale 2 as a binary covariate.

We used the *qladder* function (Stata (7)), which displays the quantiles of a transformed variable against the quantiles of a normal distribution according to the ladder powers 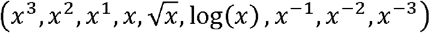 for each continuous covariate and chose the following transformations:-log_e_ (respiratory rate), log_e_ (pulse rate), log_e_ (systolic blood pressure), and log_e_ (diastolic blood pressure).

We developed all models using York Hospital (YH) data (as development dataset) and externally validated their performance on Scarborough Hospital (SH) data (as validation dataset). The hospitals are part of the same NHS Trust but are geographically separated by about 65 kilometres (40 miles).

We report discrimination and calibration statistics as performance measures for these models (8).

Discrimination relates to how well a model can separate, (or discriminate) between admissions with and without COVID-19 and is given by the area under the Receiver Operating Characteristics (ROC) curve (AUC) or c-statistic. The ROC curve is a plot of the sensitivity, (true positive rate), versus 1-specificity, (false positive rate), for consecutive predicted risks. A c-statistic of 0.5 is no better than tossing a coin, whilst a perfect model has a c-statistic of 1. In general, values less than 0.7 are considered to show poor discrimination, values of 0.7 to 0.8 can be described as reasonable, and values above 0.8 suggest good discrimination (9). The 95% confidence interval for the c-statistic was derived using DeLong’s method as implemented in the *pROC* library (10) in R (11).

Calibration is the relationship between the observed and predicted risk of COVID-19 (24) and can be readily seen on a scatter plot (y-axis observed risk, x-axis predicted risk). Perfect predictions should be on the 45° line. We internally validated and assessed the calibration for all the models using the bootstrapping approach (12, 13). The overall statistical performance was assessed using the scaled Brier score which incorporates both discrimination and calibration (8). The Brier score is the squared difference between actual outcomes and predicted risk of COVID-19, scaled by the maximum Brier score such that the scaled Brier score ranges from 0–100%. Higher values indicate superior models.

The cut-off of NEWS2 is 5 or more. This is the recommended threshold for detecting deteriorating patients and sepsis (14, 15). Therefore, we assessed the sensitivity, specificity, positive and negative predictive values and likelihood ratios for these models at NEWS2 threshold of 5+ (16). We further compared the Net Benefit for all these NEWS and NEWS2 models, which may inform the utility of the models in routine clinical practice (17). The net benefit is calculated at a particular threshold probability *p*t with total sample size *N* as follows:

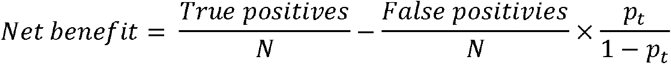

The highest net benefit has the highest clinical value.

We calculated the minimum sample size using the R package *pmsampsize* (18). We found 930 (93 events) is minimum required sample size with number of predictors =21, R2=0.182, prevalence =0.10, shrinkage>0.9, margin absolute prediction error (MAPE) = 0.05 (19). We followed the TRIPOD guidelines for reporting of model development and validation (20). We used Stata (7) for data cleaning and *R* (11) for statistical analysis.

## Results

### Cohort Characteristics

The number of non-elective discharges was 6444 over 3 months. We excluded 36 (0.6%) of admissions because the index NEWS was not recorded within ±24 hours of the admission date/time or there was missing or no recorded at all (see Table S3).

The characteristics of the admissions included in our study are shown in Table 1. Emergency admissions in the validation dataset were older than those in development dataset (69.6 years vs 67.4 years), less likely to be male (49.5% vs 51.2%), had higher index NEWS (2.8 vs 2.5) and NEWS2 (3.2 vs 2.8), higher prevalence of COVID-19 (11.0% vs 8.7%) but similar in-hospital mortality (8.4% vs 8.2%). See accompanying scatter and boxplots in Figure S1 to S4 – supplemental digital content.

We assessed the performance of index NEWS/NEWS2 models to predict the risk of COVID-19 in emergency medical admissions (see Table 2 and Figure 1). The c-statistics for predicting COVID-19 for Model M2’ is better than NEWS2 alone M0’ model in development (M0’=0.71; M1’=0.72, M2’: 0.78) and the validation dataset (M0’=0.65; M1’=0.67, M2’: 0.72). Moreover, the c-statistics for predicting COVID-19 for NEWS2 models was similar to NEWS models in both, development and validation datasets.

**Table 2:** **Performance of NEWS and NEWS2 models for predicting the risk of COVID on admission for validation dataset** ARD: absolute risk difference; AUC: Area under the curve; CIs: confidence intervals

| Model | Mean Risk Non-COVID | Mean Risk COVID | ARD | Scaled Brier Score | AUC (95% CIs) | Calibration Slope |
| --- | --- | --- | --- | --- | --- | --- |
| M0 | 0.11 | 0.14 | 0.03 | 0.01 | 0.65<br>(0.62 to 0.69) | 0.72<br>(0.53 to 0.91) |
| M1 | 0.11 | 0.14 | 0.04 | 0.02 | 0.67<br>(0.64 to 0.7) | 0.81<br>(0.63 to 0.99) |
| M2 | 0.10 | 0.18 | 0.08 | 0.05 | 0.72<br>(0.69 to 0.75) | 0.78<br>(0.65 to 0.91) |
| M0' | 0.11 | 0.14 | 0.03 | 0.00 | 0.65<br>(0.61 to 0.68) | 0.69<br>(0.51 to 0.87) |
| M1' | 0.11 | 0.14 | 0.04 | 0.01 | 0.67<br>(0.64 to 0.7) | 0.78<br>(0.60 to 0.96) |
| M2' | 0.10 | 0.19 | 0.09 | 0.06 | 0.72<br>(0.69 to 0.75) | 0.76<br>(0.64 to 0.89) |

**Figure 1.**
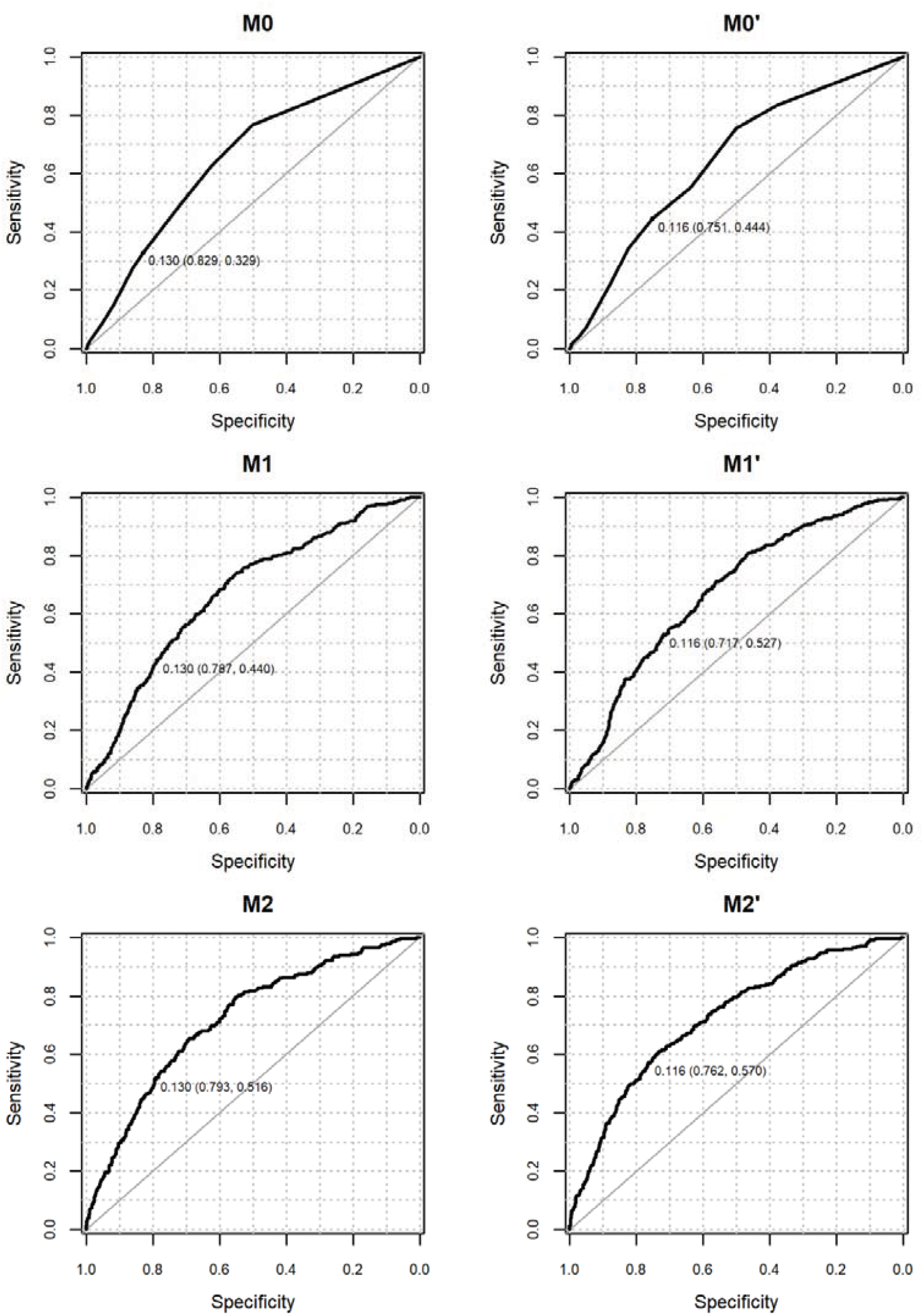
Receiver Operating Characteristic curve for NEWS (left column) and NEWS2 (right column) in predicting the risk of COVID-19 on admission for model M0/M0’, M1/M1’, and M2/M2’ in the validation dataset. Note: predicted probability at NEWS (or NEWS2) threshold ≥5 (sensitivity, specificity) is shown for all models.

Table 3 includes the sensitivity, specificity, positive and negative predictive values for NEWS and NEWS2 models for predicting COVID-19. NEWS2 models had higher sensitivity but lower specificity compared to NEWS models because the predicted probability at NEWS2≥5 (0.116) is smaller than at NEWS≥5 (0.13).

**Table 3.** **Sensitivity analysis of three models for each NEWS and NEWS2 for predicting the risk of COVID at threshold ≥5 of NEWS (predicted probability of model M0 = 0.130) and NEWS2 (predicted probability of model M0’ = 0.116) for validation dataset.** PPV=Positive Predictive Value; NPV= Negative Predictive Value; LR+=Positive Likelihood Ratio; LR-=Negative Likelihood Ratio

| Model | Number of positive cases identified by model | Sensitivity% | Specificity% | PPV | NPV | LR+ | LR- |
| --- | --- | --- | --- | --- | --- | --- | --- |
| M0 | 474 | 32.9<br>(27.4 to 38.7) | 82.9<br>(81.3 to 84.5) | 19.2<br>(15.7 to 23) | 90.9<br>(89.6 to 92.1) | 1.9<br>(1.6 to 2.3) | 0.8<br>(0.7 to 0.9) |
| M1 | 600 | 44<br>(38.1 to 50.1) | 78.7<br>(76.9 to 80.4) | 20.3<br>(17.2 to 23.8) | 91.9<br>(90.6 to 93.1) | 2.1<br>(1.8 to 2.4) | 0.7<br>(0.6 to 0.8) |
| M2 | 607 | 51.6<br>(45.6 to 57.6) | 79.3<br>(77.6 to 81) | 23.6<br>(20.2 to 27.1) | 93<br>(91.8 to 94.1) | 2.5<br>(2.2 to 2.9) | 0.6<br>(0.5 to 0.7) |
| M0' | 681 | 44.4<br>(38.5 to 50.5) | 75.1<br>(73.3 to 76.9) | 18.1<br>(15.2 to 21.2) | 91.6<br>(90.3 to 92.9) | 1.8<br>(1.5 to 2.1) | 0.7<br>(0.7 to 0.8) |
| M1' | 781 | 52.7<br>(46.6 to 58.7) | 71.7<br>(69.8 to 73.5) | 18.7<br>(16 to 21.6) | 92.5<br>(91.1 to 93.7) | 1.9<br>(1.6 to 2.1) | 0.7<br>(0.6 to 0.7) |
| M2' | 692 | 57<br>(51 to 62.9) | 76.2<br>(74.4 to 77.9) | 22.8<br>(19.8 to 26.1) | 93.5<br>(92.3 to 94.6) | 2.4<br>(2.1 to 2.7) | 0.6<br>(0.5 to 0.6) |

Model M2’ had better calibration than Model M0’ (M2’: 0.78 (95%CI 0.65 vs 0.91) vs 0.69 (95%CI 0.51 to 0.87)) (see Table 2 & S4 and Figure 1 & S7) with improved sensitivity (M2’: 57% (95%CI 51-63) vs M0’ 44% (95%CI 38-50)) and similar specificity (M2’: 76% (95%CI 74-78) vs M0’ 75% (95%CI 73-77)) for validation dataset at NEWS2≥5 (see Table 3 & S5). Internal validation of these models is shown in Figure S6. Figure 2 shows model calibration improved across the models and that models M2’ and M2 are well-calibrated.

**Figure 2.**
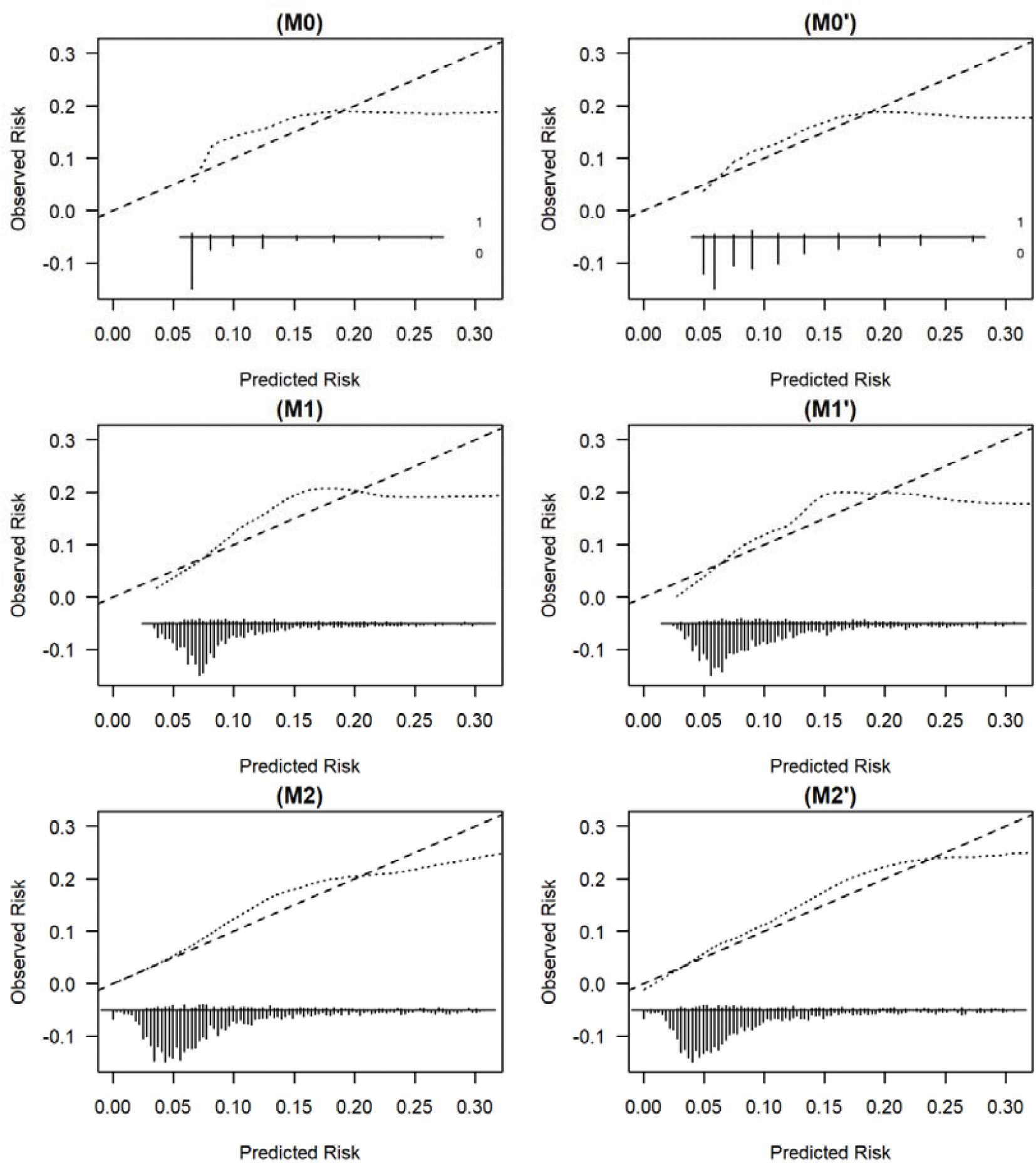
External validation of all NEWS (M0, M1, M2) and NEWS2 (M0’, M1’, M2’) models, respectively for predicting the risk of COVID-19. NB: We limit the risk of COVID-19 to 0.30 for visualisation purpose because beyond this point, we have few patients.

Model M2’/M2 had highest clinical utility than Model M0’/M0 for validation datasets (see Figure 3) and development dataset (see Figure S8). Nevertheless, NEWS/NEWS2 ≥5 is the worst performing choice compared to the cNEWS models.

**Figure 3.**
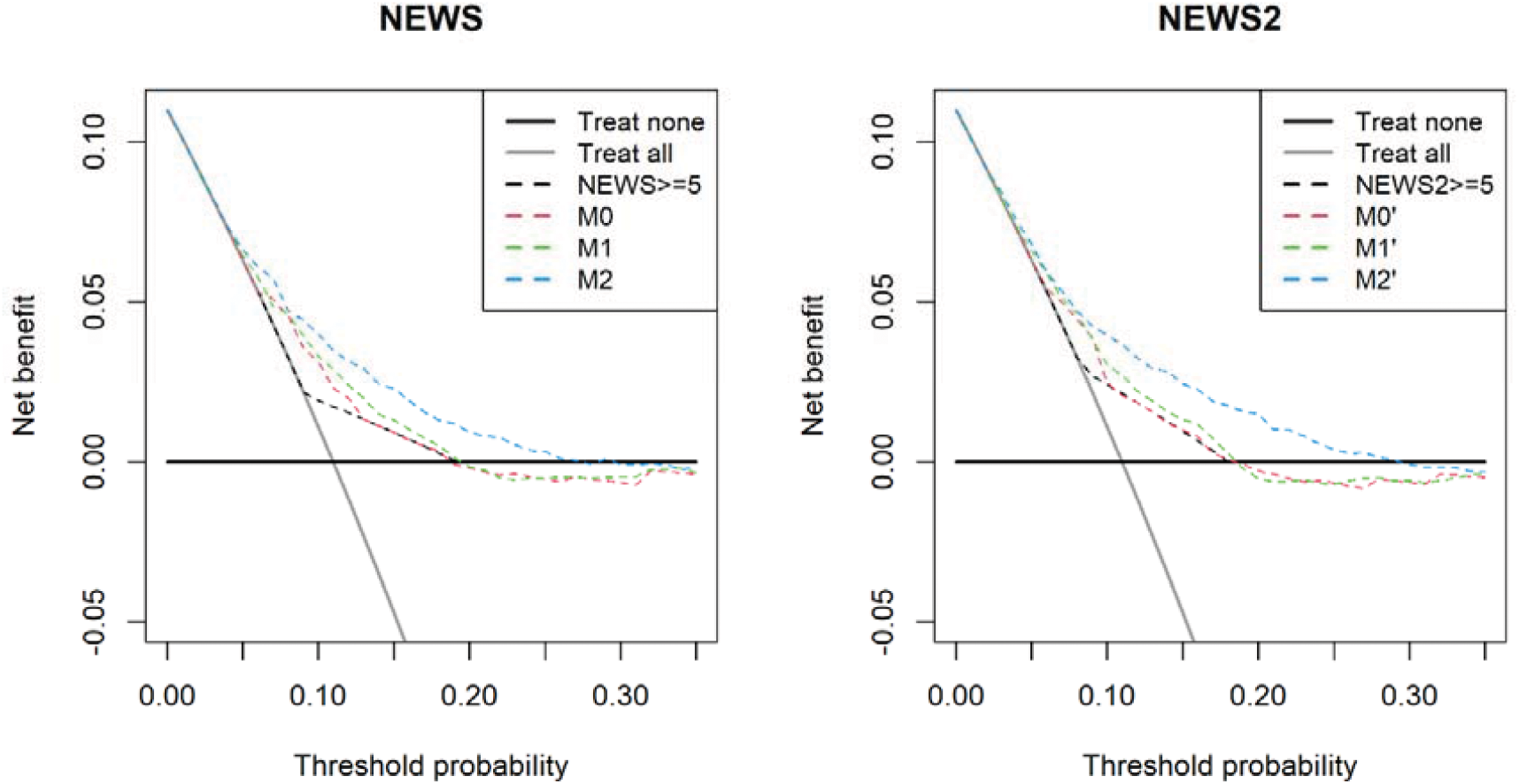
Net Benefit for model M0/M0’, M1/M1’, and M2/M2’ in predicting the risk of COVID-19 on admission in the validation dataset.

## Discussion

In this study, we developed and validated three computer-aided versions of NEWS/NEWS2 based models which incorporated progressively more information. Model M0 uses NEWS alone; Model M1 extends M0 with age and sex; Model M2 extends M1 with all the subcomponents of NEWS plus diastolic blood pressure. Equivalent models (M0’, M1’, M2’) were developed using NEWS2 (see appendix for equations and escalation policy Figure S5).

NEWS2 models were more sensitive but less specific than NEWS models. Models M2 and M2’ were the best in class, with the highest c-statistics (0.77 and 0.72 respectively). The high negative predictive value suggests models M2 and M2’ may be particularly useful in ruling out COVID-19 early in the patients unplanned admission which is clinically useful because testing for COVID-19 using viral nucleic acid testing is time consuming.

A recent systematic review identified five models to detect COVID-19 infection in symptomatic individuals with c-statistics that ranged from 0.87 to 1 (21). However, despite these high c-statistics, the review authors cautioned against the use of these models in clinical practice because of the high risk of bias and poor reporting of studies which are likely to have led to optimistic results (21).

The main advantages of our computer-aided NEWS2 models are that they are designed to incorporate data which are already available in the patient’s electronic health record and so place no additional data collection or computational burden on clinicians and they are readily automated. Nonetheless, we emphasize that our computer-aided risk scores are not designed to replace clinical judgement. They are intended and designed to support, not subvert, the clinical decision-making process and can be always overridden by clinical concern (5, 22). The working hypothesis for our computer-aided NEWS scores is that they may enhance situational awareness of COVID-19 by processing information already available without impeding the workflow of clinical staff, especially as our approach offers a faster and less expensive assessment of COVID-19 risk than current laboratory tests which may be more practical to use for large numbers of people.

There are limitations in relation to our study. We identified COVID-19 based on ICD-10 code ‘U071’ which was determined by clinical judgment and/or swab test results and so our findings are constrained by the accuracy of these methods (23, 24). We used the index NEWS or NEWS2 data in our models, which reflects the “on-admission” risk of COVID-19 of the patient. Nonetheless, vital signs are repeatedly updated for each patient according to hospital protocols. Although we developed models using one hospital data and validated into other hospital data, the extent to which changes in vital signs over time reflect changes in COVID-19 risk that need to be incorporated in our models needs further study. While most of the studies reported insufficient sample size (25), our study was sufficiently large for developing and validating relatively simple NEWS/NEWS2 based prediction models(19). Our two hospitals are part of the same NHS Trust and this may undermine the generalisability of our findings, which merit further external validation.

Furthermore, a crucial next phase of this work is to field test our models by carefully engineering then into routine clinical practice (26, 27) to see if they do support the earlier detection and care of COVID-19 in emergency medical patients without unintended adverse consequences.

## Conclusion

Model M2’ is reasonably accurate for predicting the on-admission risk of COVID-19. It may be clinically useful for an early warning system at the time of admission especially to triage large numbers of unplanned hospital admissions.

## Data Availability

Our data sharing agreement is with York hospital and does not permit us to share the data used in this paper.

## Author contributions

DR and MAM had the original idea for the work. KB, RH provided the data extracts. MF undertook the statistical analyses with support from MAM. MF, MAM, and DR wrote the first draft of the paper. DR, SI and KS provided clinical perspectives. EW provided critical guidance and support. All others contributed to the final paper and have approved the final version. DR & MF will act as study guarantors.

## Transparency declaration

The lead author (the manuscript’s guarantor) affirms that the manuscript is an honest, accurate, and transparent account of the study being reported; that no important aspects of the study have been omitted.

## Ethical Approval

This study was deemed to be exempt from ethical approval because it was classified as an evaluation. Furthermore, this study used already de-identified data from an ongoing study involving NEWS which received ethical approval from Health Research Authority (HRA) and Health and Care Research Wales (HCRW) (reference number 19/HRA/0548).

## Patient involvement

none

## Data sharing

our data sharing agreement is with York hospital and does not permit us to share the data used in this paper.

