## supplementary material for "Predictive accuracy of computer-aided versions of the on-admission National Early Warning Score in estimating the risk of COVID-19 for unplanned admission to hospital: a retrospective development and validation study"

**NEWS models (M0, M1, M2)**

**Model M0**

$$\boldsymbol{Logit}\left( \boldsymbol{COVID} \right)\boldsymbol{= -3.064+ 0.232 * NEWS}$$

**Model M1**

$$\boldsymbol{Logit}\left( \boldsymbol{COVID} \right)\boldsymbol{= -3.881+ 0.168 * Male + 0.011 * Age+ 0.224 * NEWS}$$

**Model M2**

$$\boldsymbol{Logit}\left( \boldsymbol{COVID} \right)\boldsymbol{= -5.500+ 0.126 * Male + 0.009 * Age- 0.069 * NEWS+ 1.770 * log}\left( \boldsymbol{Respiratory Rate} \right)\boldsymbol{+ 0.286 * Temperature-0.941 * log}\left( \boldsymbol{Systoloic pressure} \right)\boldsymbol{- 0.211 * log}\left( \boldsymbol{Diastolic pressure} \right)\boldsymbol{+0.300 * log}\left( \boldsymbol{Pulse rate} \right)\boldsymbol{- 0.097 * Oxygen Saturations+ 1.184 * Oxygen Supplementation- 1.462 *Pain + 0.505* Voice- 8.393}$$

**We accounted for baseline difference in risk of COVID-19 in the external validation data by adding (M0: 0.19, M1:0.18, M2:0.19) to the NEWS logit models using an iterative procedure described elsewhere^1^**

**1.	Faisal M, Howes R, Steyerberg EW, Richardson D, Mohammed MA. Using routine blood test results to predict the risk of death for emergency medical admissions to hospital: an external model validation study. QJM [Internet]. 2017 Jan 1 [cited 2017 Oct 2];110(1):27–31. Available from: https://academic.oup.com/qjmed/article-lookup/doi/10.1093/qjmed/hcw110**

**NEWS2 models (M0’, M1’, M2’)**

**Model M0’**

$$\boldsymbol{Logit}\left( \boldsymbol{COVID} \right)\boldsymbol{= -3.131+ 0.219 * NEWS}$$

**Model M1’**

$$\boldsymbol{Logit}\left( \boldsymbol{COVID} \right)\boldsymbol{= -3.946+ 0.181 * Male + 0.011 * Age+ 0.213 * NEWS}$$

**Model M2’**

$$\boldsymbol{Logit}\left( \boldsymbol{COVID} \right)\boldsymbol{=-4.147+ 0.114 * Male + 0.009* Age+0.006 * NEWS+ 1.416 * log}\left( \boldsymbol{Respiratory Rate} \right)\boldsymbol{+ 0.287 * Temperature-0.756 * log}\left( \boldsymbol{Systoloic pressure} \right)\boldsymbol{- 0.348 * log}\left( \boldsymbol{Diastolic pressure} \right)\boldsymbol{+0.167 * log}\left( \boldsymbol{Pulse rate} \right)\boldsymbol{- 0.099 * Oxygen Saturations+ 0.797 * Oxygen Supplementation- 2.185 *Pain + 0.120* Voice- 8.889 * Unconscious+0.390*Baseline Confusion+0.270*New Confusion-0.868*Scale}\boldsymbol{2+0.046*Oxygen Flow Rate}$$

**We accounted for baseline difference in risk of COVID-19 in the external validation data by adding (M0’: 0.18, M1’:0.17, M2’:0.18) to the NEWS2 logit models using an iterative procedure described elsewhere^1^**

**1.	Faisal M, Howes R, Steyerberg EW, Richardson D, Mohammed MA. Using routine blood test results to predict the risk of death for emergency medical admissions to hospital: an external model validation study. QJM [Internet]. 2017 Jan 1 [cited 2017 Oct 2];110(1):27–31. Available from: https://academic.oup.com/qjmed/article-lookup/doi/10.1093/qjmed/hcw110**

**Table S1:NEWS scoring chart**

| **Physiological Parameters** | **3** | **2** | **1** | **0** | **1** | **2** | **3** |
| --- | --- | --- | --- | --- | --- | --- | --- |
| **Respiration Rate** | **≤8** |  | **9 - 11** | **12 - 20** |  | **21 - 24** | **≥25** |
| **Oxygen Saturations** | **≤91** | **92 - 93** | **94 - 95** | **≥96** |  |  |  |
| **Any Supplemental Oxygen** |  | **Yes** |  | **No** |  |  |  |
| **Temperature** | **≤35.0** |  | **35.1 - 36.0** | **36.1 - 38.0** | **38.1 - 39.0** | **≥39.1** |  |
| **Systolic BP** | **≤90** | **91 - 100** | **101 - 110** | **111 - 219** |  |  | **≥220** |
| **Heart Rate** | **≤40** |  | **41 - 50** | **51-90** | **91 - 110** | **111 - 130** | **≥131** |
| **Level of Consciousness** |  |  |  | **Alert** |  |  | **Voice, Pain, or Unconscious** |

**Table S2: NEWS2 scoring chart**

| **Physiological Parameters** | **3** | **2** | **1** | **0** | **1** | **2** | **3** |
| --- | --- | --- | --- | --- | --- | --- | --- |
| **Respiration Rate** | **≤8** |  | **9 - 11** | **12 - 20** |  | **21 - 24** | **≥25** |
| **SpO2 Scale 1 (%)** | **≤91** | **92 - 93** | **94 - 95** | **≥96** |  |  |  |
| **SpO2 Scale 2 (%)** | **≤83** | **84 - 85** | **86 - 87** | **88 - 92**  **≥93 on Air** | **93 – 94 on oxygen** | **95 – 96 on oxygen** | **≥97 on oxygen** |
| **Oxygen Saturations** | **≤91** | **92 - 93** | **94 - 95** | **≥96** |  |  |  |
| **Air or oxygen?** |  | **Oxygen** |  | **Air** |  |  |  |
| **Temperature** | **≤35.0** |  | **35.1 - 36.0** | **36.1 - 38.0** | **38.1 - 39.0** | **≥39.1** |  |
| **Systolic BP** | **≤90** | **91 - 100** | **101 - 110** | **111 - 219** |  |  | **≥220** |
| **Heart Rate** | **≤40** |  | **41 - 50** | **51-90** | **91 - 110** | **111 - 130** | **≥131** |
| **Level of Consciousness** |  |  |  | **Alert** |  |  | **Voice, Pain, Confusion, or Unconscious** |

**The NEWS [https://www.rcplondon.ac.uk/projects/outputs/national-early-warning-score-news] is based on a scoring system in which a score is allocated to vital signs physiological measurements already undertaken when patients present to or are being monitored in hospital. A score is allocated to each as they are measured, the magnitude of the score reflecting how extreme the parameter varies from the norm. This score is then aggregated, and uplifted for people requiring oxygen.**

| **Characteristic** | **Development dataset (YH)** | **Validation dataset (SH)** | **All** |
| --- | --- | --- | --- |
|  | **N (%)** | **N (%)** | **N (%)** |
| **Total emergency medical discharges between**  **11 Mar 20 to 13 June 20** | **3952** | **2528** | **6480** |
| **Excluded: No NEWS recorded (%)** | **13 (0.3)** | **6 (0.2)** | **19 (0.3)** |
| **Excluded: First NEWS after 24 hours of admission (%)** | **15 (0.4)** | **2 (0.1)** | **17 (0.3)** |
| **Total excluded (%)** | **28 (0.7)** | **8 (0.3)** | **36 (0.6)** |
| **Total included (%)** | **3924 (99.3)** | **2520 (99.7)** | **6444 (99.4)** |

**Table S3 Number of emergency medical admissions included/excluded**

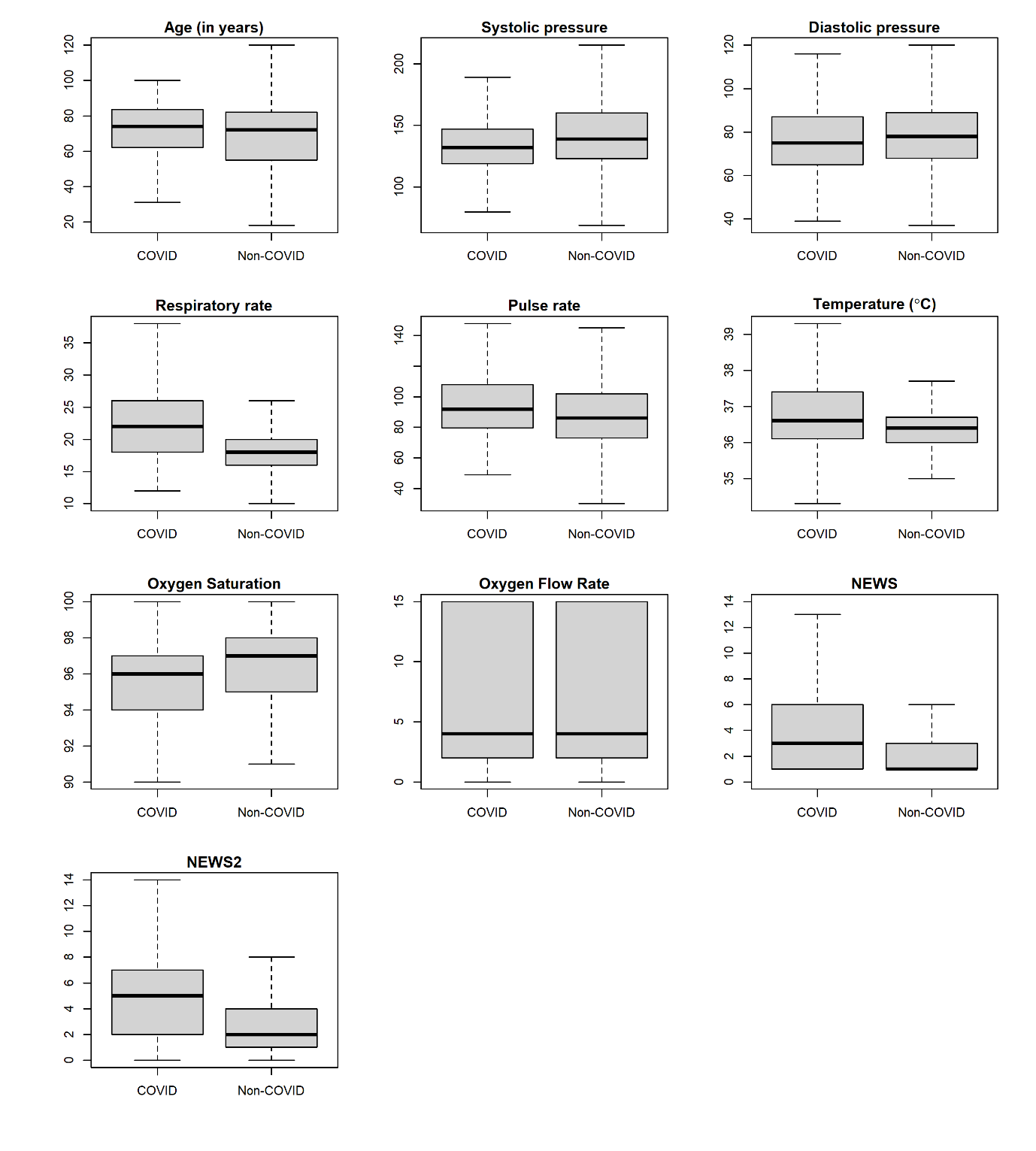

### Figure S1 Boxplot for continuous covariates without outliers with respect to COVID-19 (Yes/No) for development dataset.

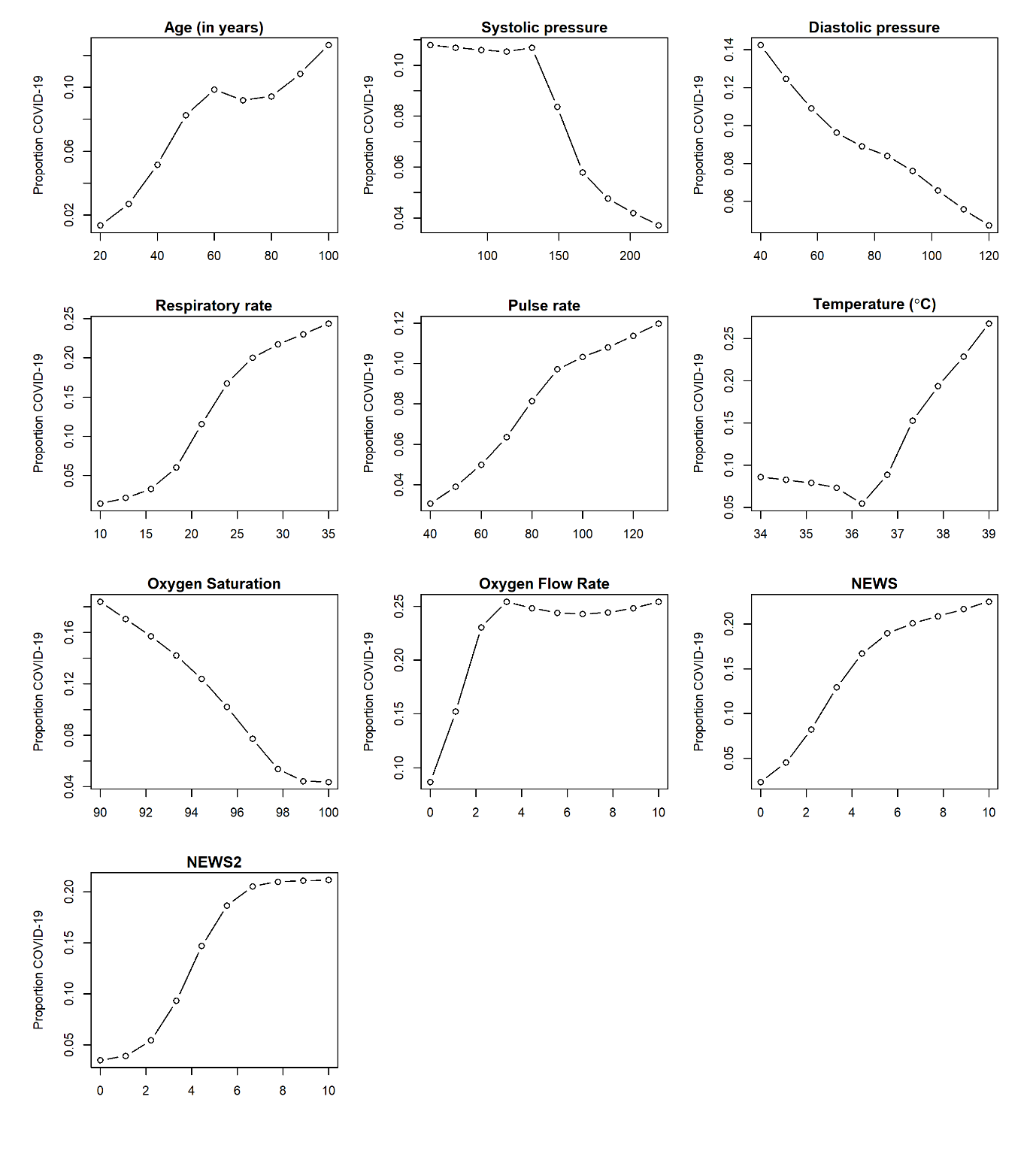

Figure S2 Scatter plots showing the observed risk of COVID-19 with continuous covariates for development dataset.

NB: y-axis range changes in each plot.

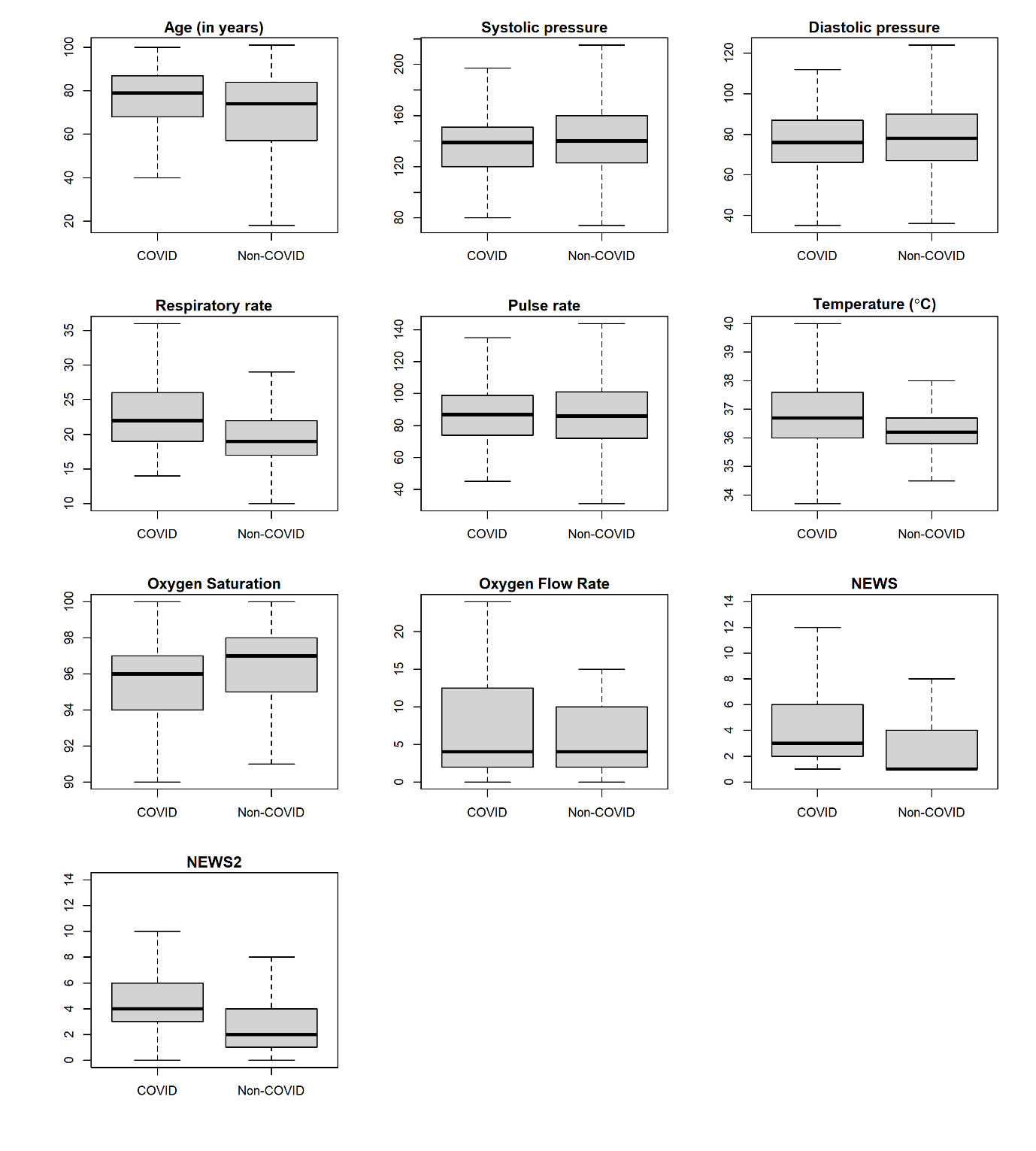
Figure S3 Boxplot for continuous covariates without outliers with respect to COVID-19 (Yes/No) for validation dataset.

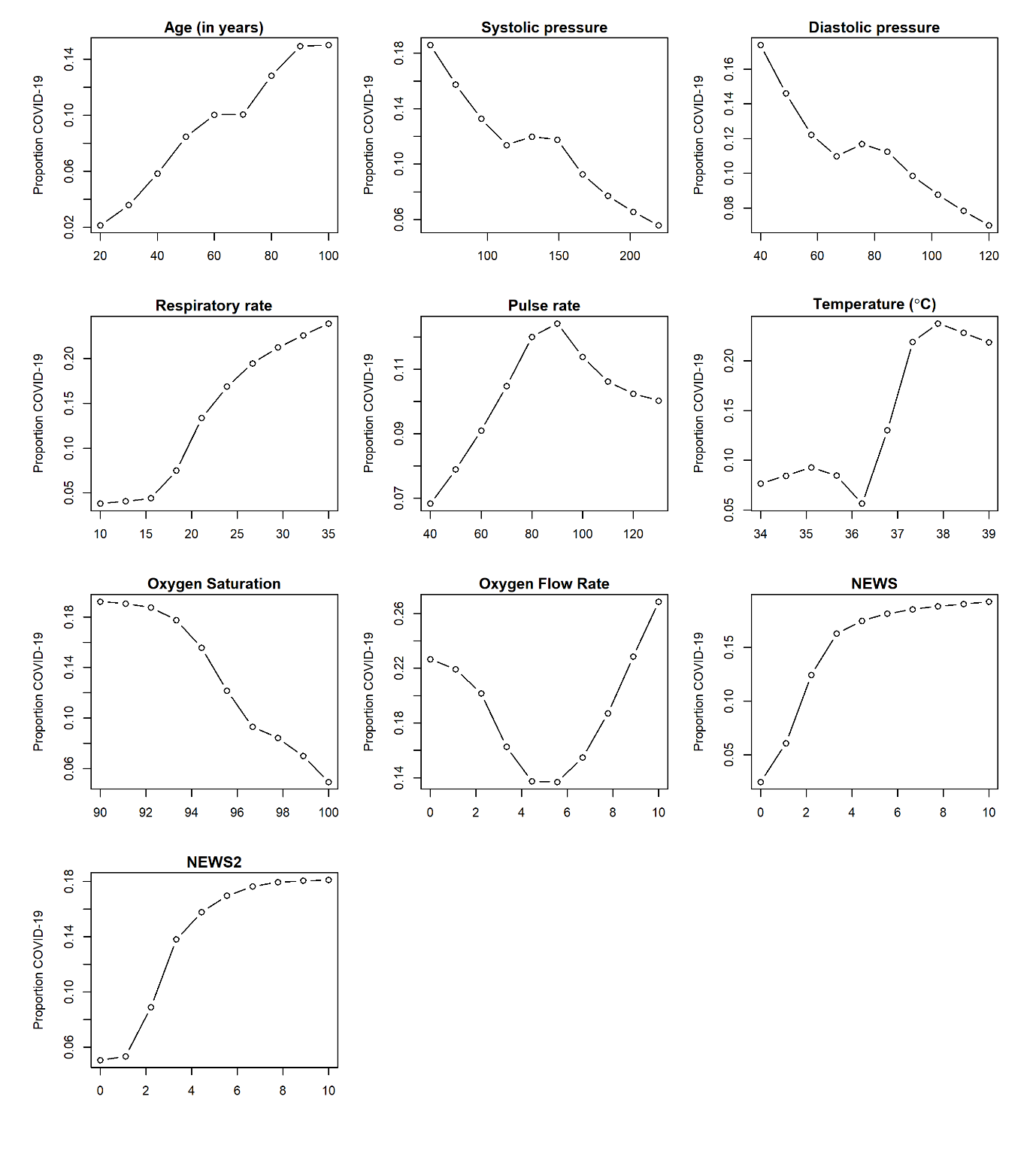
Figure S4 Scatter plots showing the observed risk of COVID-19 with continuous covariates for NH

NB: y-axis range changes in each plot.

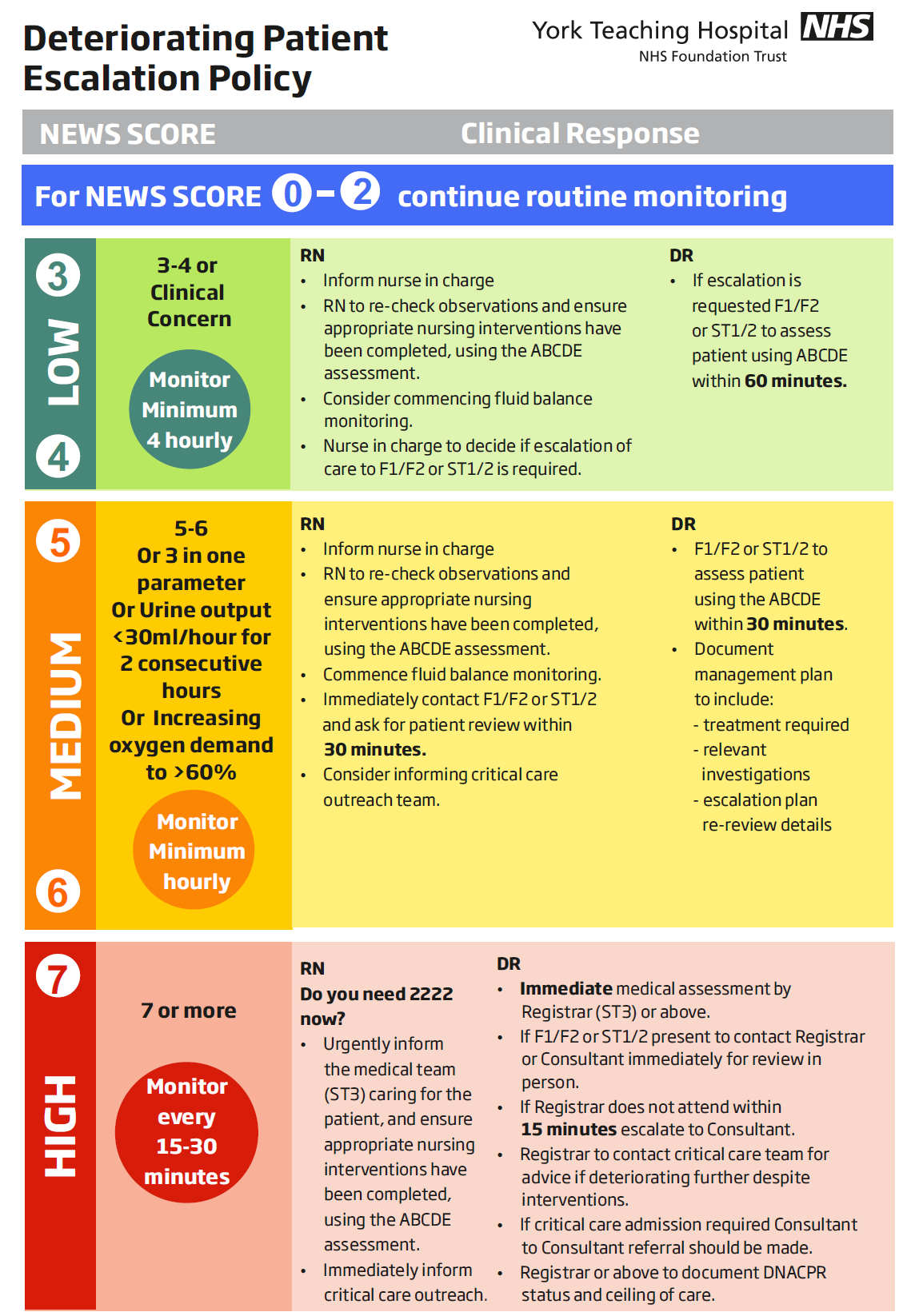

**Figure S5 Escalation policy of deteriorating patients in York Teaching Hospital NHS Foundation Trust**

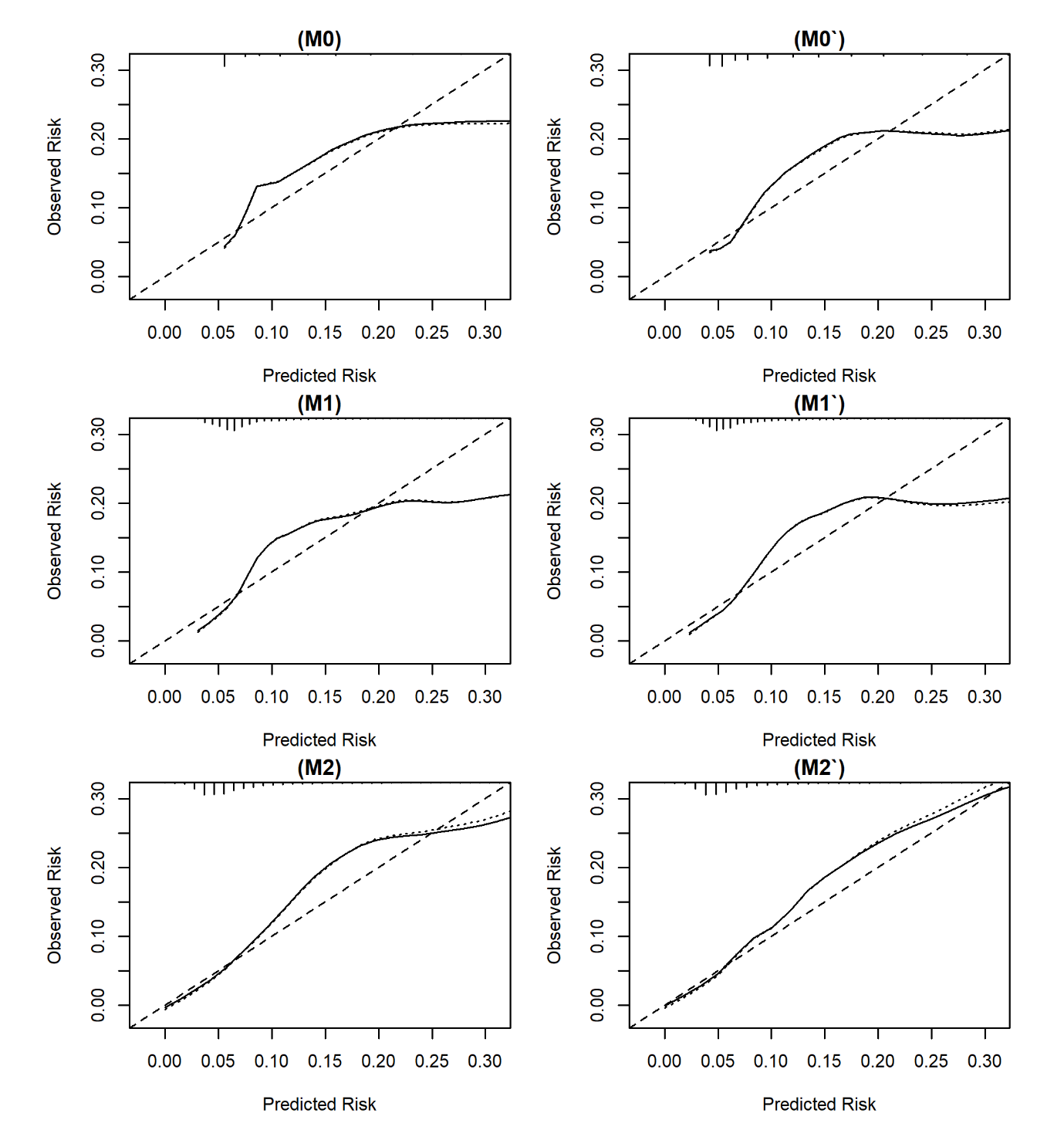

Figure S6 Calibration of NEWS (M0, M1, M2) and NEWS2 (M0’,M1’,M2’) models, respectively for predicting the risk of COVID-19

NB: We limit the risk of COVID-19 to 0.30 for visualisation purpose because beyond this point, we have few patients.

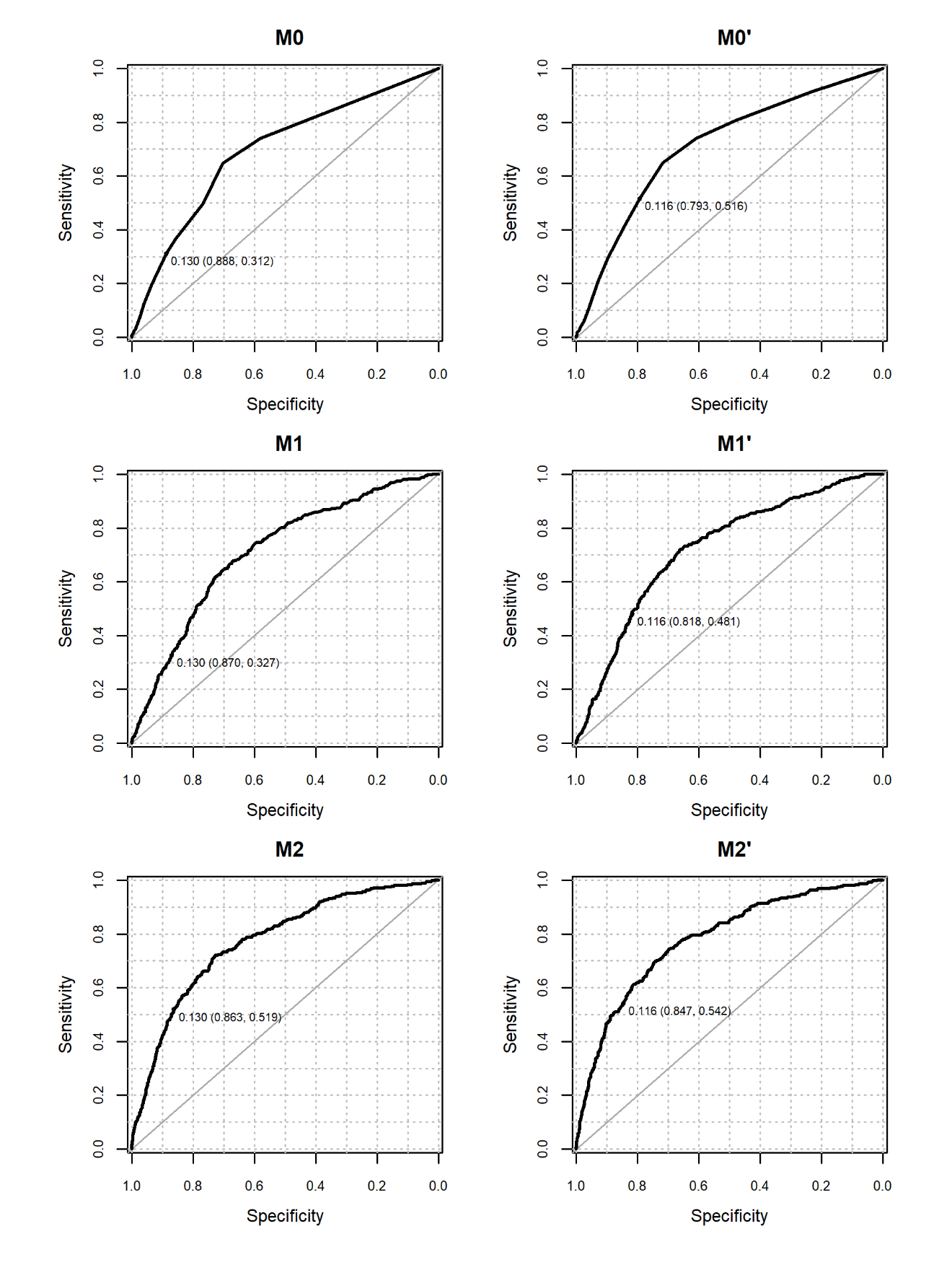

**Figure S7 Receiver Operating Characteristic curve for NEWS (left column) and NEWS2 (right column) in predicting the risk of COVID-19 in development dataset on admission for model M0/M0’, M1/M1’, and M2/M2’**

Note: predicted probability at NEWS (or NEWS2) threshold ≥5 (sensitivity, specificity) is shown for all models.

| **Model** | **Development dataset** | | | | | |
| --- | --- | --- | --- | --- | --- | --- |
|  | **Mean Risk Non-COVID** | **Mean Risk COVID** | **ARD** | **Scaled Brier Score** | **AUC (95% CIs)** | **Optimism-corrected AUC** |
| **M0** | **0.08** | **0.12** | **0.04** | **0.03** | **0.69  (0.67 to 0.72)** | **0.70** |
| **M1** | **0.08** | **0.13** | **0.04** | **0.04** | **0.71  (0.68 to 0.74)** | **0.71** |
| **M2** | **0.08** | **0.18** | **0.10** | **0.09** | **0.77  (0.75 to 0.8)** | **0.77** |
| **M0`** | **0.08** | **0.13** | **0.05** | **0.04** | **0.71  (0.68 to 0.74)** | **0.71** |
| **M1`** | **0.08** | **0.13** | **0.05** | **0.04** | **0.72  (0.7 to 0.75)** | **0.72** |
| **M2`** | **0.08** | **0.19** | **0.11** | **0.10** | **0.78  (0.75 to 0.81)** | **0.77** |

**Table S4: Performance of NEWS and NEWS2 models for predicting the risk of COVID on admission for development dataset**

**ARD: absolute risk difference; AUC: Area under the curve; CIs: confidence intervals**

| **Model** | **Number of positive cases identified by model** | **Sensitivity%** | **Specificity%** | **PPV** | **NPV** | **LR+** | **LR-** |
| --- | --- | --- | --- | --- | --- | --- | --- |
| **M0** | **508** | **31.2  (26.3 to 36.4)** | **88.8  (87.7 to 89.8)** | **21.1  (17.6 to 24.9)** | **93.1  (92.2 to 93.9)** | **2.8  (2.3 to 3.3)** | **0.8  (0.7 to 0.8)** |
| **M1** | **577** | **32.7  (27.7 to 37.9)** | **87  (85.9 to 88.1)** | **19.4  (16.3 to 22.9)** | **93.1  (92.2 to 93.9)** | **2.5  (2.1 to 3)** | **0.8  (0.7 to 0.8)** |
| **M2** | **669** | **51.9  (46.5 to 57.3)** | **86.3  (85.1 to 87.4)** | **26.6  (23.3 to 30.1)** | **94.9  (94.1 to 95.7)** | **3.8  (3.3 to 4.3)** | **0.6  (0.5 to 0.6)** |
| **M0`** | **919** | **51.6  (46.2 to 57)** | **79.3  (77.9 to 80.6)** | **19.3  (16.8 to 22)** | **94.5  (93.6 to 95.3)** | **2.5  (2.2 to 2.8)** | **0.6  (0.5 to 0.7)** |
| **M1`** | **818** | **48.1  (42.7 to 53.5)** | **81.8  (80.5 to 83)** | **20.2  (17.5 to 23.1)** | **94.3  (93.4 to 95.1)** | **2.6  (2.3 to 3)** | **0.6  (0.6 to 0.7)** |
| **M2`** | **735** | **54.2  (48.8 to 59.6)** | **84.7  (83.4 to 85.8)** | **25.3  (22.2 to 28.6)** | **95.1  (94.3 to 95.8)** | **3.5  (3.1 to 4)** | **0.5  (0.5 to 0.6)** |

**Table S5 Sensitivity analysis of three models for each NEWS and NEWS2 for predicting the risk of COVID at threshold ≥5 of NEWS (predicted probability of model M0 = 0.130) and NEWS2 (predicted probability of model M0’ = 0.116) for development dataset.**

**PPV=Positive Predictive Value; NPV= Negative Predictive Value; LR+=Positive Likelihood Ratio; LR-=Negative Likelihood Ratio**

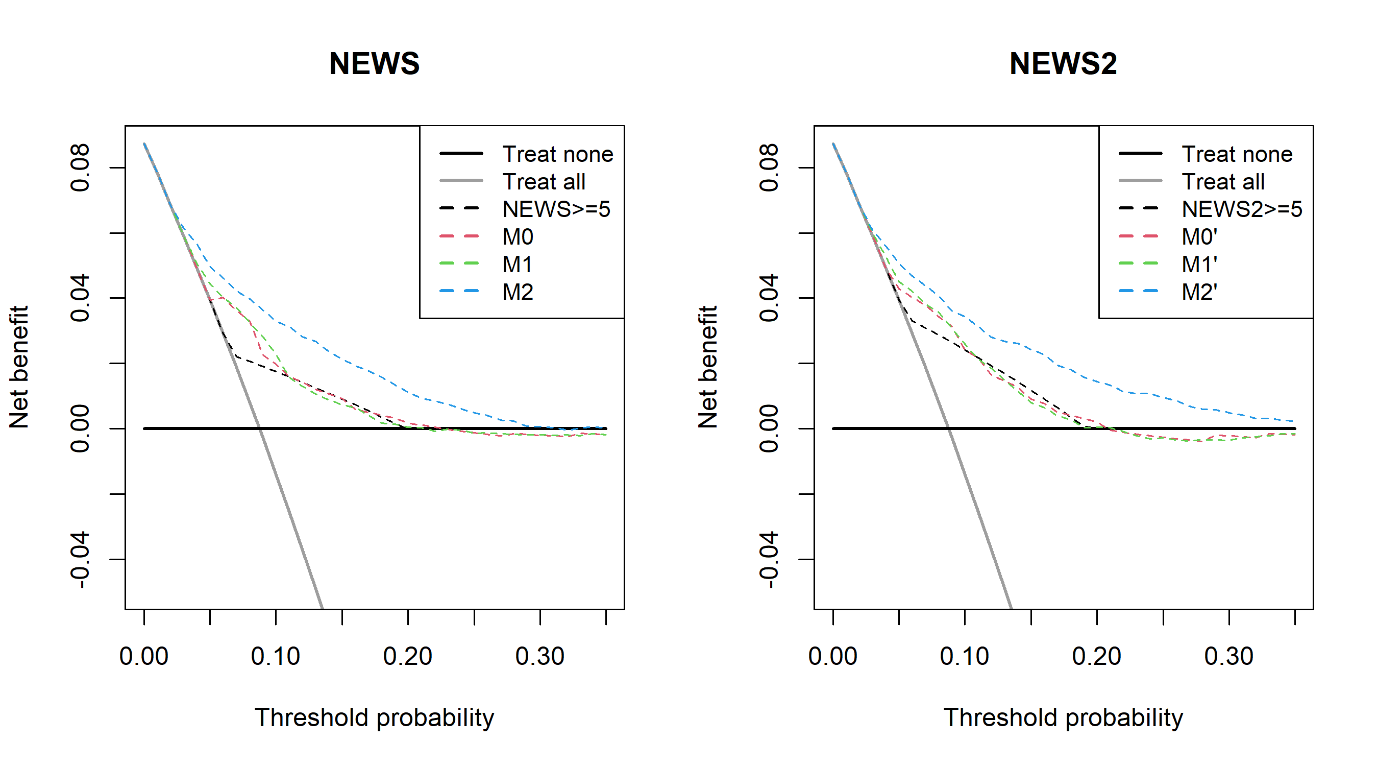

**Figure S8 Net Benefit for model M0/M0’, M1/M1’, and M2/M2’ in predicting the risk of COVID-19 on admission in the validation dataset**
